# Plasma Neurofilament Light Chain is Markedly Elevated in General Hospital Inpatients with Delirium

**DOI:** 10.64898/2026.10.18.26365854

**Authors:** Matthew JY Kang, Dhamidhu Eratne, Thomas Borchard, Kruti Patel, Charles B Malpas, Christa Dang, Meg Storer, Oneil G Bhalala, Jenny Jia Yu, Alexander F Santillo, Malcolm Forbes, Henrik Zetterberg, Sarah Farrand, Samantha M Loi, Philip B Mitchell, Malcolm Hopwood, Jeffrey Smith, Cherie Chiang, Dennis Velakoulis

## Abstract

**Objective:** To determine whether plasma neurofilament light chain (NfL) and glial fibrillary acidic protein (GFAP) are elevated in hospitalised patients with delirium relative to patients with dementia and primary psychiatric disorders (PPD).

**Methods and analysis:** This was a cross-sectional observational study conducted at the Royal Melbourne Hospital, Melbourne, Australia, between April and October 2024, with comparator samples from a research biobank. In Phase 1 (Delirium Discovery Cohort), we measured plasma NfL and GFAP using the Single molecule array (Simoa) platform in hospitalised patients with delirium and compared them with dementia and PPD groups. Biomarker concentrations were the primary outcome and were compared using general linear models adjusted for age, sex, and estimated glomerular filtration rate, with bootstrapped confidence intervals. Discriminative performance was assessed as a secondary outcome using receiver operating characteristic (ROC) curve analysis,. In Phase 2, we applied this threshold to independent dementia and PPD groups.

**Results:** In Phase 1 (n =176), inpatients with delirium showed higher concentrations of both NfL and GFAP than patients with dementia (β = 0.78; 95% CI [0.37, 1.20] and β = 0.39 [0.07, 0.73] respectively) or PPD groups (β = 1.17; [0.69, 1.66] and β = 0.83 [0.51, 1.24] respectively). ROC curve analysis for differentiating delirium from dementia and PPD groups yielded an optimal NfL threshold of 24.1 pg/mL (AUC = 0.89, 96% sensitivity, 70% specificity). In comparison, GFAP showed lower accuracy (AUC = 0.85), and combining NfL with GFAP (AUC = 0.90) did not significantly improve performance. In Phase 2 (n = 154), the NfL threshold of > 24 pg/ml maintained strong performance (AUC = 0.95, 96% sensitivity, and 77% specificity).

**Conclusions:** Plasma NfL shows promise as a biomarker for delirium in hospitalised patients, distinguishing it with high sensitivity from patients with dementia and PPD. Larger multicentre studies are needed to refine cut-offs, evaluate temporal dynamics, and determine integration into acute hospital delirium pathways.

**What is already known on this topic:** Delirium is highly prevalent in general hospitals but remains frequently underdiagnosed. Currently, no validated blood-based biomarker exists to reliably assist in clinical identification.

**What this study adds:** Plasma neurofilament light chain (NfL) is substantially elevated in general hospital inpatients with delirium compared to those with dementia or primary psychiatric disorders. This includes patients with delirium superimposed on dementia, yielding a discriminative profile well-suited for ruling out the diagnosis.

**How this study might affect research, practice or policy:** These findings support the prospective evaluation of plasma NfL as an objective adjunct to clinical bedside assessments. This biomarker could significantly aid in evaluating patients with acute confusion, particularly when hypoactive presentations make clinical diagnosis challenging.

## 1. INTRODUCTION

Delirium is an acute neuropsychiatric syndrome characterised by fluctuating confusion and inattention,^1^ which is highly prevalent among hospitalised older adults and linked to serious outcomes including prolonged hospitalisation, cognitive decline, and death.^2,3^ However, delirium is frequently missed or misattributed as other conditions including dementia and psychiatric disorders.^4,5^ This is because delirium is associated with numerous neuropsychiatric symptoms such as hallucinations, delusions, agitation, apathy, and altered psychomotor activity, and these symptoms overlap with dementia and psychiatric conditions. When clinically suspected, misdiagnosis rates range from 10-60%.^6–8^ Moreover, underdiagnosis is a major problem, with previous literature identifying that 86% of delirium cases meeting DSM-5 criteria were not formally recorded as discharge diagnoses.^5^ Missed or misdiagnosed delirium prevents appropriate investigation of precipitating causes, leads to inappropriate treatment, and results in inadequate follow-up.^9^ A recent review of delirium did not identify any blood biomarker that can reliably assist in its identification, which currently relies almost solely on clinical assessment.^10^ An objective biomarker could aid screening and diagnosis, and could significantly reduce the health and social costs related to undiagnosed delirium.^11–13^

The development of ultra-sensitive immunoassays has enabled measurement of brain-derived proteins that cross the blood-brain barrier. Two biomarkers have shown promise in delirium. Neurofilament light chain protein (NfL), a marker of neuronal injury, predicted those who developed post-operative delirium^14,15^ and was associated with longer duration of delirium and coma in intensive care unit (ICU) settings.^16–18^ Krogseth et al. found that an acute delirium episode led to NfL elevations persisting up to six months.^19^ Glial fibrillary acidic protein (GFAP), a marker of astrogliosis, is elevated in postoperative and ICU delirium.^15,20,21^ However, existing studies have focused almost exclusively on perioperative or ICU delirium.

Critically, no study has assessed NfL or GFAP among general hospital inpatients with delirium, despite this being the most common clinical setting where delirium occurs.^22,23^ Delirium outside perioperative and intensive care contexts is unique in several important ways. It arises from diverse precipitating factors (infections, metabolic disturbances, polypharmacy) rather than surgical trauma. Delirium occurs in a more heterogeneous patient population with varied comorbidities, and often presents without the controlled perioperative monitoring that enables early detection in surgical cohorts. Additionally, no prior work has directly compared biomarker levels across delirium, dementia, and psychiatric disorders in the same dataset, despite these being the most common conditions clinicians must distinguish when evaluating acute confusion or altered mental status in hospitalised older adults.^2^

Interpretation is complicated by the fact that NfL is also elevated in dementia, reflecting chronic neurodegeneration,^24–26^ and appears mildly elevated in some psychiatric disorders.^26–29^ Furthermore, NfL levels increase with age,^30^ adding complexity when interpreting results in older adults. This makes it difficult to discern the underlying cause of elevated NfL levels in older adults with cognitive impairment. The critical question is whether delirium produces a sufficiently distinct elevation in NfL or GFAP to enable detection even when baseline levels are elevated due to age, dementia, or psychiatric conditions. If so, blood biomarkers could aid in identifying patients who require deeper clinical assessment, investigation of precipitating causes, and delirium-specific management. These knowledge gaps highlight the need to evaluate the diagnostic performance of NfL and GFAP in real-world hospital cohorts of delirium, dementia, and primary psychiatric disorders.

The primary aim of this study (Aim 1) was to determine whether plasma NfL and GFAP concentrations are elevated in hospitalised patients with delirium compared to those with dementia or a primary psychiatric disorder (PPD). We hypothesised that both biomarkers would be higher in the delirium group, regardless of whether the delirium is isolated or superimposed on dementia. As a secondary aim (Aim 2), we evaluated the diagnostic performance of plasma NfL and GFAP in separating delirium from its comparator groups (dementia and PPD). As exploratory analyses, we aimed to assess biomarker heterogeneity within the delirium group by comparing delirium-only and delirium with comorbid dementia subgroups against the dementia group (Aim 3a), and associations between biomarkers and delirium-specific clinical variables including severity scores and clinical features (Aim 3b).

## 2. METHODS

This study is reported in accordance with the STROBE statement for observational studies. The primary outcomes were the adjusted group differences in plasma NfL and GFAP concentrations. Secondary and exploratory analyses evaluated the discriminative performance of these biomarkers. We employed a two-phase design (Figure 1):

**Figure 1.**
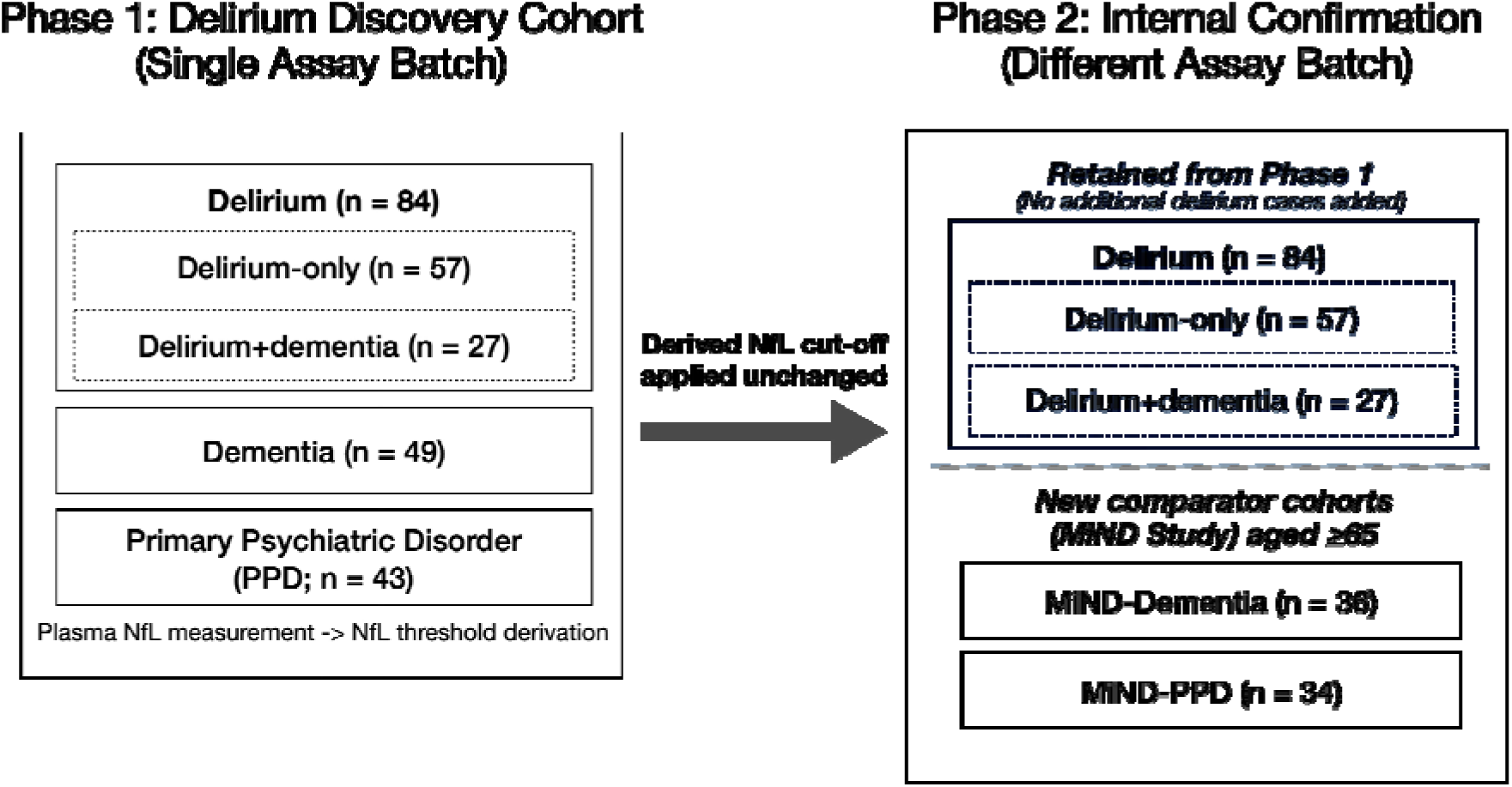
Two-phase derivation and internal confirmation of a plasma neurofilament light (NfL) threshold. Phase 1 derived an NfL cut-off in a Delirium Discovery Cohort processed in a single assay batch. The derived cut-off was applied unchanged in Phase 2 by retaining the original delirium samples and pairing them with independent dementia and primary psychiatric disorder comparator cohorts from The MiND (Markers in Neuropsychiatric Disorders) Study.

**Phase 1 (Delirium Discovery Cohort)** compared patients with delirium (with and without comorbid dementia) against dementia and PPD groups. This phase established the primary group differences and derived the candidate NfL threshold.

**Phase 2 (Threshold Application)** retained the Phase 1 delirium cohort but substituted independent dementia and PPD comparators who were older and tested in separate assay batches. This allowed us to assess the threshold’s robustness across age differences.

### 2.1 Delirium Discovery Cohort (Phase 1)

Patients with delirium in this study were admitted to the Royal Melbourne Hospital (a tertiary hospital with medical, surgical, and psychiatric units) between April and October 2024. Patients were prospectively identified through screening of emergency department lists and delirium clinical nurse specialist referrals. The delirium clinical nurse specialist receives referrals from the hospital for inpatients with suspected delirium to assist with diagnosis and management. Blood samples from patients with delirium were collected during acute delirium, corresponding to the time of clinical nurse specialist assessment or peak delirium severity. Delirium diagnoses were confirmed by a consultant neuropsychiatrist through review of the electronic medical record, based on DSM-5-TR criteria supported by routinely collected 4 Assessment Test (4AT)^31^ and Delirium Observation Screening Scale (DOSS) assessments.

Patients with dementia were similarly identified from the screening of emergency department lists and delirium clinical nurse specialist referrals. Patients with a primary psychiatric disorder (PPD) were identified using the emergency department and acute inpatient psychiatry unit lists as previously described.^29^ Remnant blood samples were collected during their index admission. Because dementia (n = 16) and PPD (n = 22) groups were under-represented, these groups were supplemented with BeYOND participants (33 dementia and 15 PPD).^33,34^ BeYOND participants were patients admitted to the specialist neuropsychiatry department of the same hospital. Dementia and PPD diagnoses were verified through consultant neuropsychiatrist review of clinical records.

We obtained clinical data including age, sex, as well as routine pathology test results where available, including estimated glomerular filtration rate (eGFR), albumin levels, HbA1C, and the anticholinergic burden as calculated by the ACB Calculator.^35^ The ACB Calculator provides a quick sum of a patient’s anticholinergic burden from their medications based on two validated scales.^36,37^ For the delirium group, we also collected 4AT and DOSS scores.

Inclusion criteria for Phase 1 were age ≥50 years and availability of remnant plasma samples for biomarker analysis. The age criterion was selected to ensure age-comparable comparison groups, given the known influence of age on NfL levels.^30^ Patients were not excluded based on medical comorbidities, including conditions known to elevate NfL (such as renal impairment, recent stroke, or central nervous system infections), as the study aimed to evaluate biomarker performance in a representative general hospital population. Two patients initially identified in the PPD group were excluded due to acute alcohol intoxication at the time of blood sampling, as this may acutely confound biomarker levels and obscure the underlying psychiatric diagnosis.^38,39^

### 2.2 Phase 2 comparator cohorts for internal testing

Phase 2 addressed a key limitation of Phase 1, which was the substantial age difference between delirium and comparators (Table 1). Independent testing cohorts were drawn from The Markers in Neuropsychiatric Disorders Study (The MiND Study),^24,40^ and were restricted to participants aged ≥65 years to better approximate the age distribution of the delirium cohort. These cohorts comprised dementia and primary psychiatric disorder (PPD) groups. The MiND-Dementia group (n = 36 after applying age restriction) comprised a range of neurodegenerative conditions, including Alzheimer’s disease, synucleinopathies, behavioural variant frontotemporal dementia, Parkinson’s plus syndromes, Huntington’s disease, and mixed or vascular dementias. The MiND-PPD group (n = 34) consisted of individuals admitted at psychiatric inpatient hospitals for their primary psychiatric disorder. The Phase 1 delirium group was retained, so that the Phase 1 NfL threshold could be applied against new comparator groups.

**Table 1.** Participant characteristics across Phase 1 and Phase 2. Expressed as Mean (SD) and n (%). DOSS = Delirium Observation Screening Scale; eGFR (ml/min/1.73 m²) = estimated glomerular filtration rate; GFAP = glial fibrillary acidic protein; NfL = neurofilament light chain; PPD = primary psychiatric disorder. Values are mean (SD) for continuous variables and n (%) for categorical variables. Phase 2 used the same delirium cohort from Phase 1 (N=84) paired with age-restricted (≥65 years) independent comparator cohorts from the MiND Study. Weight was not available for Phase 2 MiND cohorts or BeYOND cohorts.

|  | Phase 1: Delirium Discovery Cohort |  |  | Phase 2: MiND |  |
| --- | --- | --- | --- | --- | --- |
| Characteristic | Delirium (+/- Dementia)<br>N = 84 | Dementia Only<br>N = 49 | PPD Only<br>N = 43 | Dementia<br>N = 36 | PPD<br>N = 34 |
| Age (years) | 82.6 (11.1) | 63.7 (8.5) | 61.2 (10.5) | 70.7 (4.8) | 73.5 (6.4) |
| Sex |  |  |  |  |  |
| F | 39 (48%) | 22 (45%) | 22 (51%) | 11 (31%) | 17 (50%) |
| M | 43 (52%) | 27 (55%) | 21 (49%) | 25 (69%) | 17 (50%) |
| eGFR | 61.2 (24.7) | 80.3 (13.5) | 80.7 (16.1) | 76.5 (11.3)<br>N = 29 | 68.7 (13.2)<br>N = 33 |
| Weight (kg) | 69.7 (17.1)<br>N = 82 | 73.1 (18.7)<br>N = 33 | 74.8 (18.3)<br>N = 24 | 84.3 (19.0)<br>N = 27 | 79.8 (18.6)<br>N = 32 |
| NfL (pg/mL) | 113 (135) | 33 (36) | 25 (45) | 18 (15) | 23 (9) |
| GFAP (pg/mL) | 426 (537) | 168 (132) | 94 (58) | 190 (113) | 185 (85) |

### 2.3 Sample analysis

All blood samples were stored at −80°C. Phase 1 samples (Delirium Discovery Cohort including BeYOND participants) were collected in lithium heparin tubes, while Phase 2 samples (MiND-Dementia and MiND-PPD) were collected in EDTA tubes as previously described.^24^ Plasma NfL and GFAP were measured using N2PB kits on a Quanterix Simoa HD-X analyser. The sample analysts were blinded to clinical data and diagnoses. All Phase 1 samples were analysed in a single assay batch. Phase 2 samples were analysed in separate batches. To account for batch and tube-type differences across Phase 1 and Phase 2, we used the harmonised biomarker values derived from a large reference cohort.

### 2.4 Statistical analysis

All analyses were conducted in R (version 4.5.1).

**Primary analyses (Aim 1):** General linear models (GLMs) were fitted separately for log-transformed plasma NfL and GFAP concentrations, with diagnostic group as the primary predictor and age, sex, and eGFR as covariates in line with recent best-practice recommendations for blood biomarker studies.^41^ The delirium group served as the reference category. Model coefficients and 95% confidence intervals were estimated using non-parametric bootstrapping with 10,000 replicates. Model coefficients (β) are reported as positive values indicating the magnitude by which delirium exceeds comparison groups (e.g., β = 0.70 indicates delirium had 0.70 units higher log-transformed NfL than the comparison group). Supplementary tables show the original model output with negative coefficients for comparison groups. Raw biomarker values are reported in descriptive summaries and figures.

**Diagnostic performance (Aim 2):** Receiver operating characteristic (ROC) curve analyses were performed using unadjusted biomarker values to investigate diagnostic utility between delirium and comparator groups. Area under the curve (AUC) values were compared between models using NfL alone, GFAP alone, and NfL+GFAP combined (using the sum of standardised values). Bootstrapping was used to compare ROC curves. Optimal cutoffs were selected based on Youden’s J statistic from Phase 1 data, which maximises the sum of sensitivity and specificity. This threshold was then used in Phase 2. Additional diagnostic test parameters were computed: positive and negative likelihood ratios, positive and negative predictive values, and overall accuracy.

**Sensitivity analyses (Aim 2):** To evaluate the robustness of the primary ROC findings, we conducted additional analyses. First, we examined whether incorporating age and eGFR as covariates in ROC models improved discriminative accuracy, given the substantial age difference between diagnostic groups and the known association between both variables and NfL. Second, we restricted ROC analyses to delirium versus dementia (excluding PPD) to confirm threshold robustness for this clinically relevant distinction. Third, analyses were repeated in hospital-recruited participants only to assess performance in a more homogeneous sample. Fourth, we derived a specificity-weighted Youden threshold as an alternative cutoff.

**Exploratory analyses (Aim 3):** We explored biomarker heterogeneity by separating delirium into delirium-only and delirium plus dementia subgroups, compared against dementia, excluding PPD as the contrast of interest was delirium vs dementia. Within the delirium group, we examined associations between biomarkers and clinical variables (4AT score, DOSS score, hallucinations, CRP, albumin, HbA1c, anticholinergic burden, admission duration) using bootstrapped GLMs adjusted for age, sex, and eGFR with false discovery rate (FDR) correction applied across the seven predictors tested for each biomarker.

### 2.5 Ethical considerations

Ethics approval was obtained from the Melbourne Health Human Research Ethics Committee (2016.038, 2017.090, 2018.371, 2020.142, HREC/65332/mh-2020). Patients and the public were not involved in the design, conduct, or reporting of this study.

## 3. RESULTS

### 3.1 Participant characteristics

Phase 1 (Delirium Discovery Cohort) comprised 176 participants: 84 with delirium (57 without dementia, 27 with comorbid dementia), 49 with dementia, and 43 with primary psychiatric disorder (PPD). Phase 2 paired the same 84 patients with delirium against 70 independent comparators from the MiND Study, comprising 36 individuals with dementia and 34 with PPD.

Participants with delirium were older than both Phase 1 comparison groups, with a mean age of 82.6 years (SD 11.1) against 63.7 years (SD 8.5) in the dementia group and 61.2 years (SD 10.5) in the PPD group. Within the delirium group, those with comorbid dementia were older than those without (85.4 [SD 7.7] versus 81.3 years [SD 12.2]). The Phase 2 comparators were closer in age to the delirium group than in Phase 1, at 70.7 years (SD 4.8) for dementia and 73.5 years (SD 6.4) for PPD.

Renal function also differed across groups. Mean eGFR was 61.2 mL/min/1.73m2 (SD 24.7) among patients with delirium, compared with 80.3 (13.5) in the dementia group and 80.7 (16.1) in the PPD group in Phase 1 comparators, and 76.5 (11.3) in the dementia group to 68.7 (13.2) in the PPD group in Phase 2. Full characteristics of both phases are presented in Table 1, with the Phase 1 cohort disaggregated by BeYOND and hospital groups in Supplementary Table.

### 3.2 Plasma NfL and GFAP across diagnostic groups in the Delirium Discovery Cohort (Aim 1)

Plasma NfL was significantly elevated in patients with delirium compared to dementia (β = 0.78, 95% CI [0.37, 1.20]; p < .001) and in PPD (β = 1.17, 95% CI [0.71, 1.66]; p < 0.001) adjusting for age, sex, and eGFR. Back-transformed, these correspond to concentrations approximately 2.2-fold and 3.2-fold higher in delirium than in dementia and PPD respectively (Figure 2). Renal function was independently associated with NfL (β = −0.01, 95% CI [−0.02, −0.01]; p < 0.001), while age (β = 0.01, 95% CI [-0.00, 0.02]; p = 0.124) and sex (β = 0.11, 95% CI [-0.14, 0.36]; p = 0.391) were not (S1 Table).

**Figure 2.**
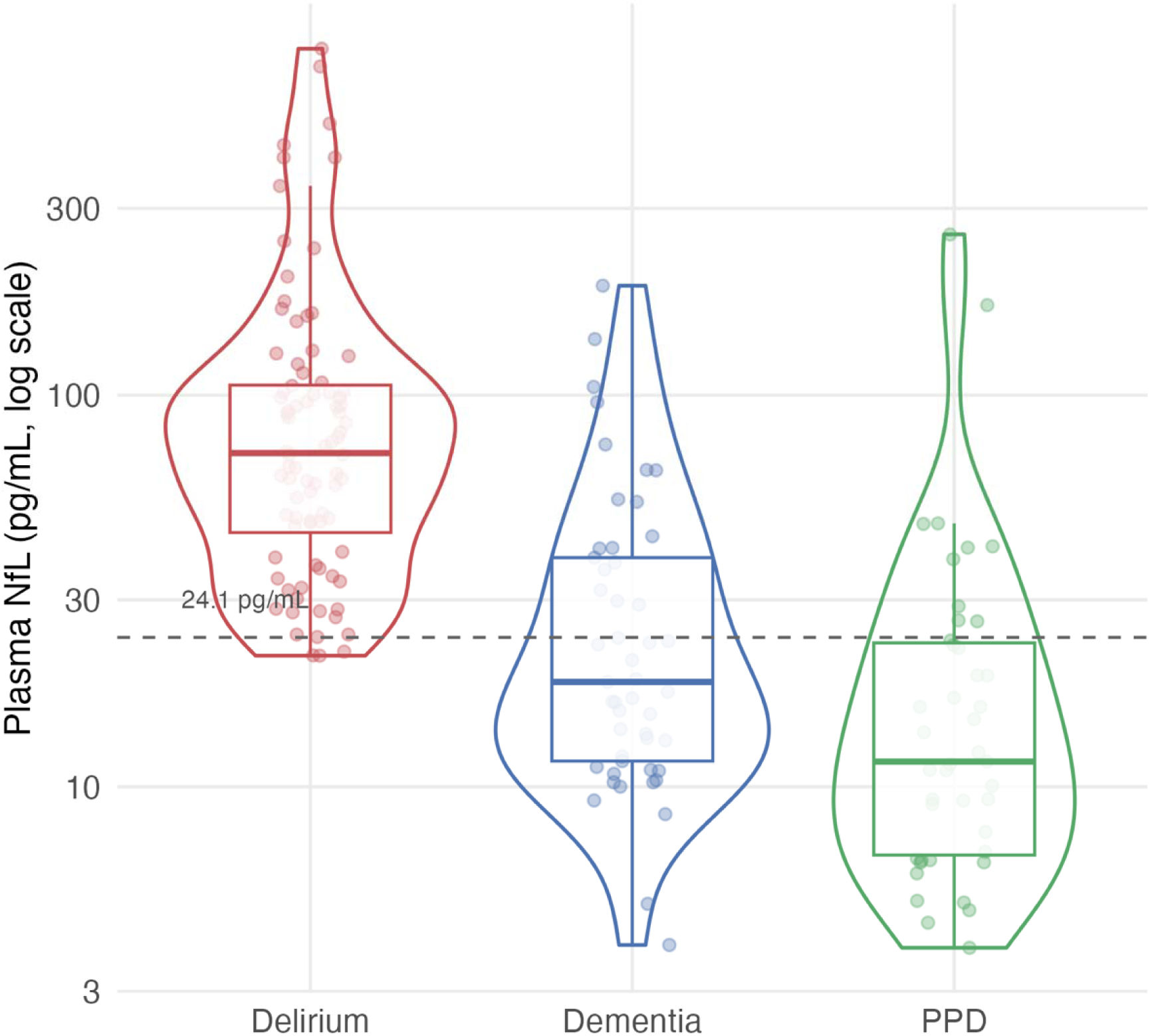
Distribution of plasma neurofilament light chain (NfL) concentrations across diagnostic groups. Violin plot with individual data points overlaid. NfL levels were highest in participants with delirium, followed by dementia only, and lowest in primary psychiatric disorders (PPD). The p values were derived from bootstrapped GLM adjusting for age, sex and eGFR. The full GLM model is available in the supplementary material. NfL = Neurofilament light chain, PPD = primary psychiatric disorders.

Plasma GFAP followed the same pattern with smaller separation (Figure 3). After adjustment for age, sex, and eGFR, GFAP higher in delirium compared to dementia (β = 0.39, 95% CI [0.07, 0.73]; p = 0.019) and in PPD (β = 0.83, 95% CI [0.51, 1.24]; p < 0.001). GFAP was associated with age (β = 0.02, 95% CI [0.01, 0.03]; p < 0.001) and sex (β = −0.26, 95% CI [−0.47, −0.05]; p = 0.014) but not eGFR (β = 0, 95% CI [-0.01, 0.00]; p = 0.142). The full model is available in the supplementary files (S2 Table).

**Figure 3.**
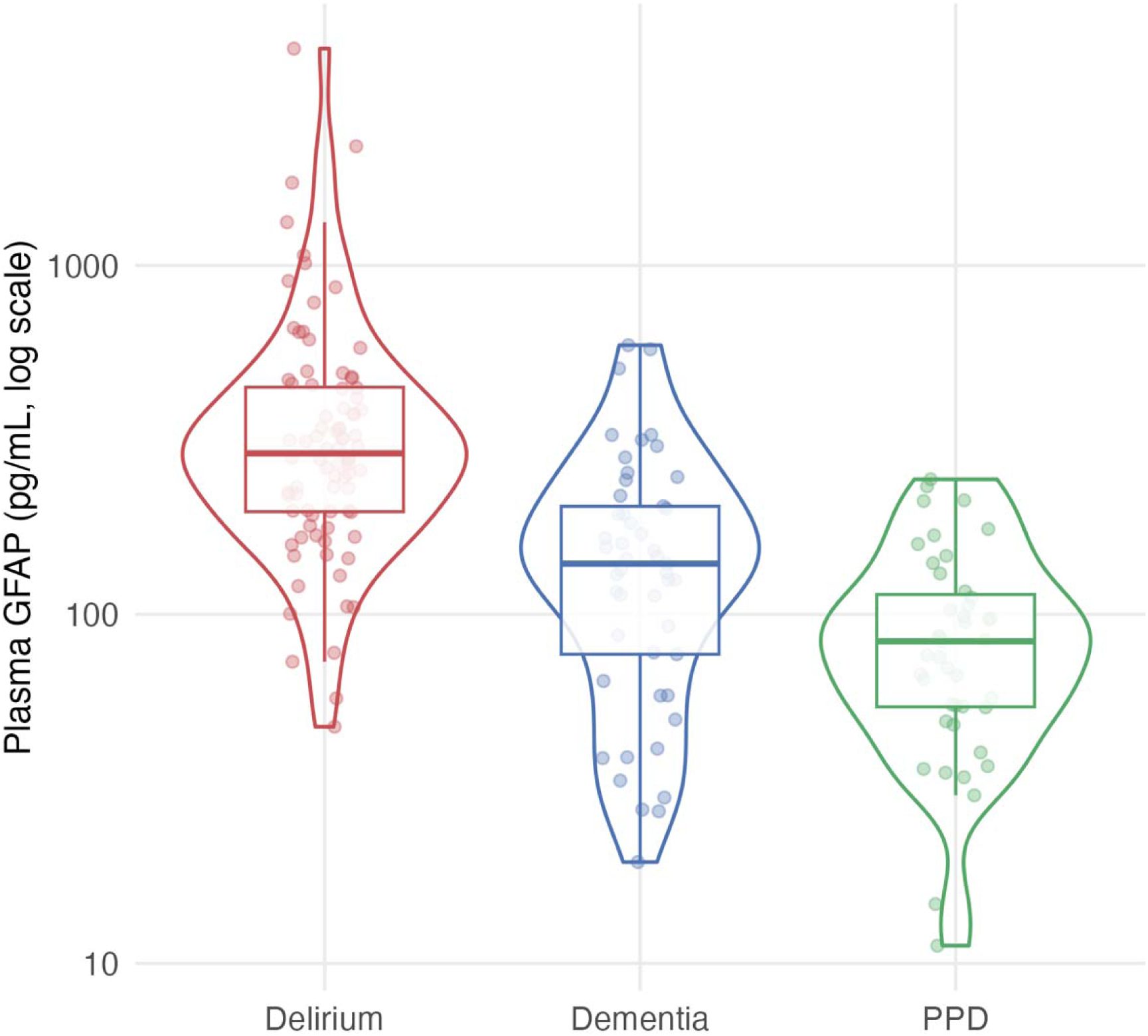
Distribution of plasma glial fibrillary acidic protein (GFAP) concentrations across diagnostic groups. Violin plot with individual data points overlaid, with GFAP levels were highest in participants with delirium (with and without comorbid dementia), intermediate in dementia, and lowest in primary psychiatric disorders (PPD). The p values were derived from bootstrapped GLM adjusting for age, sex and eGFR. The full GLM model is available in the supplementary material. GFAP = glial fibrillary acidic protein, PPD = primary psychiatric disorder

### 3.3 Discrimination between delirium and comparison groups (Aim 2)

In the receiver operating characteristic (ROC) analysis plasma NfL (Figure 4) discriminated delirium from dementia and PPD combined with an area under the curve (AUC) of 0.89 (95% CI 0.84, 0.94). Applying Youden’s index, the derived threshold of 24.1 pg/mL yielded 96% sensitivity, 70% specificity, 74% PPV, 95% NPV, and 82% overall accuracy.

**Figure 4.**
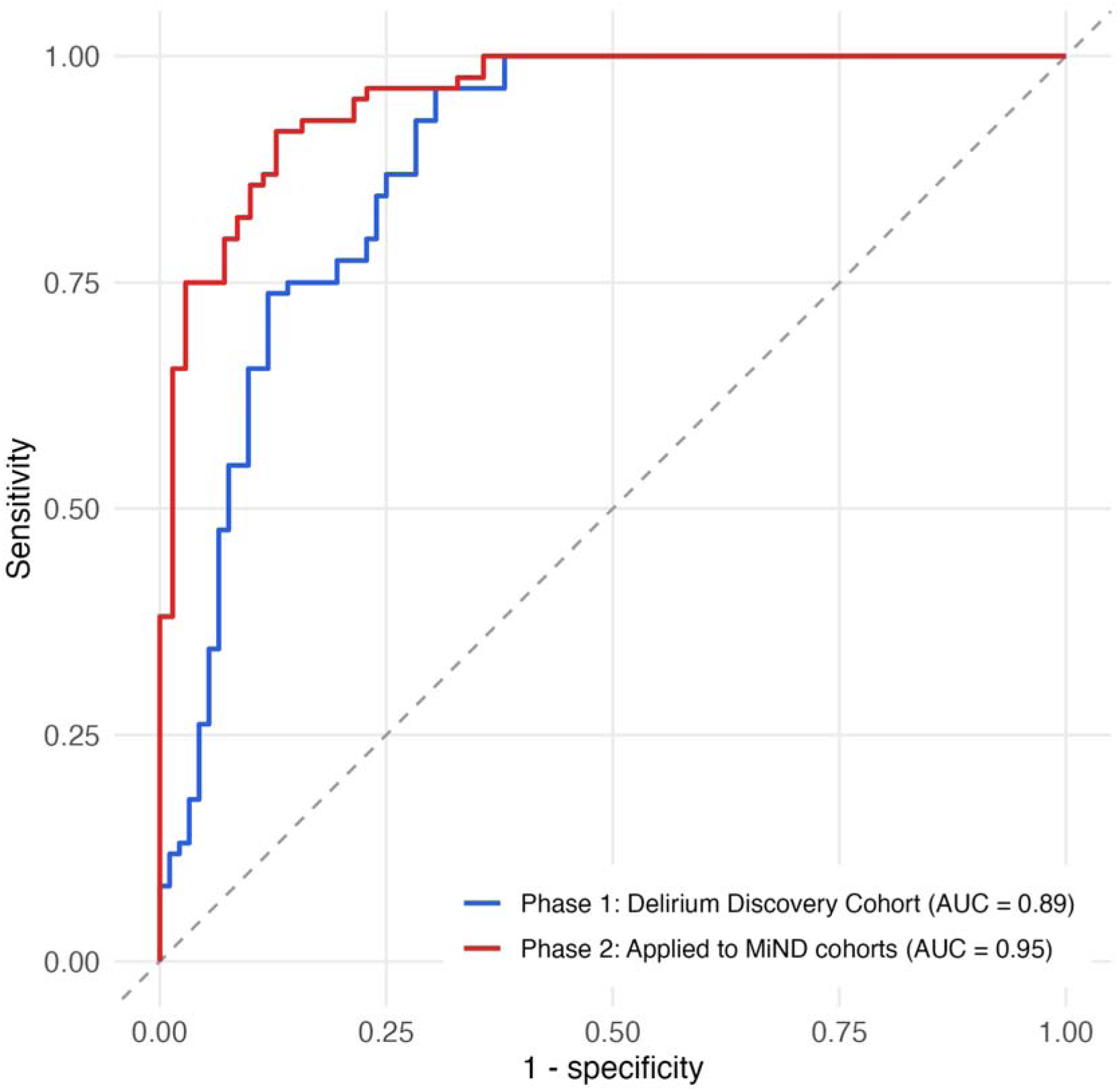
ROC curves for NfL discriminating delirium from comparator groups in the discovery and internal testing cohorts. Receiver operating characteristic (ROC) curves comparing the performance of plasma neurofilament light (NfL) in distinguishing delirium from dementia and PPD. In the first phase of the Delirium Discovery Cohort (blue), the area under the curve (AUC) was 0.89. Testing the threshold against an independent cohort of MiND-dementia and MiND-PPD groups (red) demonstrated excellent discriminative accuracy (AUC = 0.95), supporting cross-batch robustness of the derived NfL cut-off.

Plasma GFAP discriminated less well (AUC = 0.85, 95% CI [0.80, 0.91]), although the difference between markers was not significant (p = 0.303). Combining both markers did not significantly improve discrimination over NfL alone (AUC = 0.90, 95% CI [0.85, 0.95]; p = 0.260). Subsequent analyses therefore focus on NfL.

Applying the Phase 1 threshold of 24.1 pg/mL to Phase 2, plasma NfL separated delirium from the MiND comparators well overall (AUC = 0.96; 95% CI: 0.93, 0.98), yielding 96% sensitivity, 77% specificity, 83% PPV, 95% NPV, and 88% accuracy (Figure 4, Table 2).

**Table 2.** Characteristics within the delirium group with comparison to the dementia group of the Delirium Discovery Cohort including BeYOND. Values are mean (SD) or n (%). CRP, albumin, 4AT and DOSS scores not collected for dementia group. DOSS = Delirium Observation Screening Scale; eGFR (ml/min/1.73 m²) = estimated glomerular filtration rate; GFAP = glial fibrillary acidic protein; NfL = neurofilament light chain; PPD = primary psychiatric disorder.

| Characteristic | Delirium-only<br>N = 57 | Delirium + dementia<br>N = 27 | Dementia-only<br>N = 49 |
| --- | --- | --- | --- |
| Age (years) | 81.3 (12.2) | 85.4 (7.7) | 63.7 (8.5) |
| Sex: Female | 26 (46%) | 13 (48%) | 22 (45%) |
| eGFR | 62.0 (25.0) | 59.5 (24.7) | 80.3 (13.5)<br>N = 47 |
| Weight (kg) | 72.9 (18.4) | 62.6 (11.2)<br>N = 25 | 73.1 (18.7)<br>N = 33 |
| Time since admission (days) | 7.4 (8.6) | 6.3 (7.3) | 12.1 (17.5)<br>N = 16 |
| NfL (pg/mL) | 129.5 (155.1) | 78.4 (67.3) | 33 (36) |
| GFAP (pg/mL) | 375.8 (381.6) | 532.7 (768.7) | 167.6 (131.7) |
| CRP | 55.9 (61.5)<br>N = 51 | 42.3 (54.3)<br>N = 25 | 3.7 (4.2)<br>N = 10 |
| Albumin (mmol/L) | 31.9 (6.0)<br>N = 56 | 34.2 (5.2) | 37.8 (4.6)<br>N = 16 |
| 4AT score | 5.5 (3.5) | 6.5 (2.6) | N/A |
| DOSS score | 5.7 (3.3)<br>N = 39 | 6.55 (2.61)<br>N = 22 | N/A |

We examined a specificity-favoured threshold using a weighted Youden criterion (weights 0.3/0.7), which found an alternative threshold of 47.1pg/mL which had lower sensitivity (74%) and higher specificity (88%) and lower accuracy (81%) within the Phase 1 Delirium Discovery Cohort. When this cut-off was applied to the Phase 2 MiND comparator cohorts, the performance was still strong though slightly worse in accuracy (74% sensitivity, 97% specificity, 97% PPV, 76% NPV and 84% accuracy).

### 3.4 Sensitivity analyses

**Age adjustment:** Participants with delirium were approximately 20 years older than the Phase 1 comparator groups, and age was associated with plasma NfL levels (p = 0.002). We therefore compared logistic models containing each biomarker alone against models additionally containing age. For plasma NfL, adding age did not significantly improve diagnostic performance (AUC = 0.93, 95% CI [0.90, 0.97]) compared to NfL alone (p = 0.126). Adding age to plasma GFAP similarly did not reach significant difference (p = 0.157)

**Renal function:** Reduced eGFR was consistently and independently associated with higher NfL across all models. Despite this, incorporating it as an additional predictor in the ROC analysis did not improve classification accuracy (AUC remained 0.89). Full covariate adjustment (age + eGFR) for NfL yielded an AUC of 0.93 (95% CI 0.90, 0.97), demonstrating minimal additional benefit beyond age and NfL adjustments.

**Delirium-dementia distinction:** Excluding PPD, plasma NfL distinguished delirium from dementia with an AUC of 0.85 (95% CI 0.78, 0.93). At 24.1 pg/mL, sensitivity was 96% and specificity 63%.

**Hospital-based cohort:** When restricting to hospital-based participants only (n=126: 84 delirium, 16 dementia, 26 PPD), the Youden-optimal NfL threshold was 47.1 pg/mL (AUC = 0.82; 95% CI: 0.73, 0.90), yielding 74% sensitivity, 79% specificity, and 75% accuracy. When threshold of 24.1 pg/mL was applied to the hospital-only cohort, it maintained high sensitivity (96%) but with reduced specificity (52%) and comparable overall accuracy (82%).

### 3.5 Biomarker levels within the delirium and dementia groups (Aim 3a)

Plasma NfL concentration was significantly higher in those with delirium-only, compared to those with delirium and comorbid dementia (β = 0.36; 95% CI: 0.05, 0.66) and dementia-only (β = 1.05, 95% CI: 0.61, 1.52). Complete model parameters are provided in Supplementary Table S3. For GFAP, there was no significant difference in levels between delirium-only and delirium with comorbid dementia. The delirium-only group had significantly higher GFAP levels than the dementia-only group (β = 0.38, 95% CI: 0.01, 0.81; Supplementary Table S4).

### 3.6 Exploratory associations within the delirium (Aim 3b)

Two associations with NfL survived correction. Lower serum albumin was associated with higher NfL (β = 0.05; 95% CI 0.02, 0.08), and a longer interval between admission with blood sampling was associated with higher NfL (β = 0.02; 95% CI 0.01, 0.06). For GFAP, lower HbA1C was associated with higher concentrations (β = 0.10; 95% CI 0.03, 0.16).

In the remainder of the models, neither NfL nor GFAP showed a significant association with delirium factors, including the 4AT total score, DOSS average score, the presence of hallucinations (as part of the DOSS assessment), CRP levels, and anticholinergic burden.

## 4. DISCUSSION

This is the study to compared plasma NfL and GFAP levels in patients with acute delirium in the general hospital setting compared to clinically relevant comparators, dementia and primary psychiatric disorder (PPD) groups. As hypothesised, we found that both NfL and GFAP were markedly elevated in hospitalised patients with delirium. After adjustment, plasma NfL concentrations were approximately 2.2-fold higher in delirium than in dementia and 3.2-fold higher than in PPD, with plasma GFAP showing the same direction of effect at smaller magnitude. Elevation was evident in patients with delirium superimposed on dementia.

Our results build on prior reports of elevated NfL in delirium by directly comparing NfL levels in patients with delirium to patients with dementia in general hospital inpatients. General hospital inpatient settings are the most common setting in which delirium occurs yet the least studied with respect to biomarkers.^14–16,19^ Clinically, this distinction is crucial as delirium often occurs on a background of dementia, yet the management of these conditions differs substantially. The observed NfL elevation in delirium, even when dementia is present, suggests that the acute neuronal injury secondary to delirium is additional to the NfL elevation expected from baseline neurodegeneration. This aligns with epidemiological evidence linking delirium with accelerated cognitive decline and incident dementia,^3,4^ and supports the view that delirium represents a biologically injurious process rather than a transient or purely functional disturbance.^42^ Given that NfL elevations may persist up to six months following a delirium episode, future studies should examine the temporal dynamics of biomarker resolution through serial sampling. We did not collect data on prior delirium history in our comparator groups, which could have influenced baseline biomarker levels. Longitudinal studies are needed to determine whether NfL and GFAP follow similar temporal patterns, how quickly these markers normalise after delirium resolves, whether persistent elevation predicts long-term cognitive outcomes, and whether biomarker kinetics differ between delirium-only and delirium plus dementia presentations.

The marked NfL elevations observed during delirium suggest acute neuroaxonal injury, exceeding levels typically seen in chronic neurodegeneration.^24,26,43^ Multiple mechanisms may contribute to elevated NfL in delirium, including direct neuronal damage, altered blood-brain barrier permeability allowing increased NfL release into circulation, and acute CNS processes such as inflammation or metabolic disturbances. While our study did not systematically exclude patients with conditions such as stroke or CNS infections that independently elevate NfL, this heterogeneity reflects the real-world hospital population where delirium commonly occurs. Recognising delirium early facilitates systematic prevention, early detection, and timely management, particularly given established associations between delirium and subsequent cognitive decline.^44–46^

In exploratory analyses, participants with delirium-only had higher plasma NfL concentrations than those with delirium with comorbid dementia. This may indicate that acute neuroaxonal stress contributes more prominently to the biomarker signal than chronic neurodegenerative burden. One hypothesis is that in the absence of underlying dementia, a greater physiological insult may be required to precipitate delirium, leading to more pronounced neuronal injury or blood-brain barrier dysfunction. However, the relatively modest effect size and wide variability (SDs: 158 and 67 pg/mL, respectively) suggest NfL should not be used clinically to differentiate delirium-only from delirium with comorbid dementia. Collateral history, longitudinal cognitive information, structural neuroimaging, and other biomarkers (such as p-tau and amyloid markers) remain essential for identifying underlying dementia. These exploratory observations require replication in larger samples.

In contrast to NfL, plasma GFAP showed weaker discriminative performance. GFAP levels were higher in patients with delirium than in patients with primary psychiatric disorders, and in contrast to studies focused exclusively on post-operative delirium, we observed higher GFAP concentrations in our hospitalised cohort with delirium than in dementia.^14,47^ This difference may reflect variations in the underlying pathophysiology of general medical delirium compared with post-operative delirium, including the possibility of greater astroglial activation and neuroinflammation in medically precipitated delirium. Alternatively, it may indicate that post-operative delirium represents a distinct clinical context in which astroglial injury is less pronounced. Given these discrepancies, further work is needed to clarify how GFAP behaves across delirium in various clinical settings.

If our findings are further validated, NfL could serve as a rapid adjunctive tool in hospital delirium care pathways. Importantly, plasma biomarkers could be measured at hospital admission or even in community settings by general practitioners, enabling earlier delirium detection before formal clinical assessment. In older patients with acute confusion, markedly elevated NfL might strengthen suspicion of delirium, prompting structured assessments such as 4AT and DOSS. Beyond diagnostic utility, NfL shows promise for prognostication. Very high NfL levels have been associated with mortality across various conditions,^19^ and some of the highest NfL values in our cohort occurred in patients who died during admission although this information was not captured systematically. Future studies should explore whether serial NfL measurements can track delirium resolution, predict long-term cognitive outcomes, or monitor response to interventions. Such longitudinal data could inform risk stratification and guide follow-up care planning.

The Youden-optimized threshold of 24.1 pg/mL prioritised sensitivity (96%) over specificity (63%), which is appropriate for a screening context where missing delirium has serious consequences. However, examining the ROC curve reveals an alternative cutoff at approximately 47.1 pg/mL that achieves higher specificity (88%) while maintaining reasonable sensitivity (74%). This higher threshold may be more suitable in clinical scenarios where false positive results would trigger resource-intensive investigations or prolong unnecessary hospitalisation. The choice between these cutoffs should be guided by the clinical context, available resources, and consequences of missed versus over-diagnosed cases. Moreover, considering the context of the NfL level is crucial for accurate interpretation given that plasma NfL is a non-specific marker of neuronal injury and is elevated in other conditions including traumatic brain injury and stroke,^48,49^ This is where serial NfL levels may assist, as persistent or rising NfL should prompt broader evaluation for ongoing neurological injury including, but not limited to, delirium.

Several limitations of our study require consideration. Firstly, the samples were collected opportunistically rather than as part of a systematised protocol, thus we did not standardise the timing of biomarker sampling in relation to delirium onset or resolution, which may have attenuated associations with delirium severity and duration. The study lacked an acutely unwell but non-delirious medical control group, which would have strengthened interpretation of biomarker specificity. We did not exclude patients with medical conditions known to elevate NfL (such as recent stroke or CNS infections), as our aim was to evaluate NfL in a real-world hospital population where such comorbidities are common. However, this heterogeneity may have contributed to the wide range of NfL values observed in the delirium cohort and could complicate interpretation in individual patients. Our cohort generally had preserved renal function, thus larger studies including individuals with moderate to severe renal impairment are needed to determine whether incorporating eGFR meaningfully improves clinical interpretation of NfL.

Furthermore, delirium and comparator samples were drawn from different cohorts and processed across multiple assay batches. We were also unable to externally validate the diagnostic performance due to the lack of another independent delirium cohort. We were unable to adjust for other known influences on NfL, including body mass index, because of missing data. Larger studies with more complete phenotyping, including body-mass index, delirium subtype (i.e. hypoactive vs hyperactive), and delirium causes are needed to replicate and refine these findings.

## 5. CONCLUSION

Plasma NfL levels are a promising biomarker for the detection of delirium, with strong discriminatory accuracy against dementia and psychiatric disorders. In particular, high NfL levels in a person with known dementia or PPD should alert the clinician to the possibility of concurrent delirium in the appropriate clinical setting. Our findings highlight the neuropathological burden of delirium and show that NfL levels could support earlier detection and risk stratification. The widespread availability of NfL testing in the general hospital setting could result in significant diagnostic, economic and social benefits. Future studies should replicate these findings in larger cohorts, refine cut-offs, and test serial and multi-marker panels. Trials examining whether interventions that attenuate NfL elevations improve outcomes would provide important translational evidence. Ultimately, biomarker-informed approaches could improve delirium care in older adults by enabling earlier recognition, more accurate diagnosis, and a deeper understanding of its neuropathology.

## Data availability statement

De-identified data may be made available to researchers following approval of a written proposal by the study investigators and the relevant Human Research Ethics Committees, and completion of a data sharing agreement. Requests should be directed to the corresponding author.

## Contribution statement

MK and DE conceived and designed the study. MK, TB, and DE collected clinical data and samples. HZ and KB performed biomarker assays. MK and CBM performed the statistical analysis. MK drafted the manuscript. All authors contributed to interpretation of the data, critically revised the manuscript for important intellectual content, and approved the final version.

## Data Availability

Data produced in the study are available upon reasonable request to the authors.

## Supplementary sTable

| Characteristics | <u>**BeYOND**</u> |  | <u>**RMH Pathology**</u> |  |
| --- | --- | --- | --- | --- |
|  | **Dementia**<br>**N = 33** | **PPD**<br>**N = 17** | **Dementia**<br>**N = 16** | **PPD**<br>**N = 26** |
| Age | 60.2 (5.1) | 58.4 (4.5) | 70.9 (9.7) | 63.1 (12.7) |
| Sex | NA | NA | NA | NA |
| F | 12 (36%) | 8 (47%) | 10 (63%) | 14 (54%) |
| M | 21 (64%) | 9 (53%) | 6 (38%) | 12 (46%) |
| eGFR<br>(mL/min/1.73m<sup>2</sup>) | 85.3 (6.9)<br>N = 31 | 84.8 (8.7) | 70.4 (17.6) | 78.1 (19.1) |
| Weight (kg) | 79.2 (17.5)<br>N = 18 | 80.6 (18.0)<br>N = 11 | 65.9 (18.0)<br>N = 15 | 71.3 (18.2)<br>N = 15 |
| NfL (pg/mL) | 26.1 (37.6) | 7.9 (3.8) | 46.6 (28.3) | 36.7 (54.9) |
| GFAP (pg/mL) | 134.6 (90.3) | 53.5 (27.9) | 235.6 (175.3) | 119.7 (58.3) |

**S1.** NfL GLM (Reference = delirium) within Delirium Discovery Cohort.

S1. NfL GLM (Reference = delirium) within Delirium Discovery Cohort
| **Parameter** | **Coefficient [95% CI]** | **p** |
| --- | --- | --- |
| Intercept | 4.24 [2.96, 5.56] | <0.001 |
| Diagnosis: Dementia (vs Delirium) | -0.78 [-1.19, -0.37] | <0.001 |
| Diagnosis: PPD (vs Delirium) | -1.17 [-1.66, -0.71] | <0.001 |
| Age | 0.01 [0, 0.02] | 0.124 |
| Male sex | 0.11 [-0.14, 0.36] | 0.377 |
| eGFR | -0.01 [-0.02, -0.01] | <0.001 |

Note: The delirium group served as the reference category. Model coefficients are shown with their original negative signs (indicating lower NfL in comparison groups). In the main text, these are reported as positive values for clarity (e.g., $\beta = -0.70$ for dementia is reported as $\beta = 0.70$ indicating delirium had higher NfL).

**S2.**
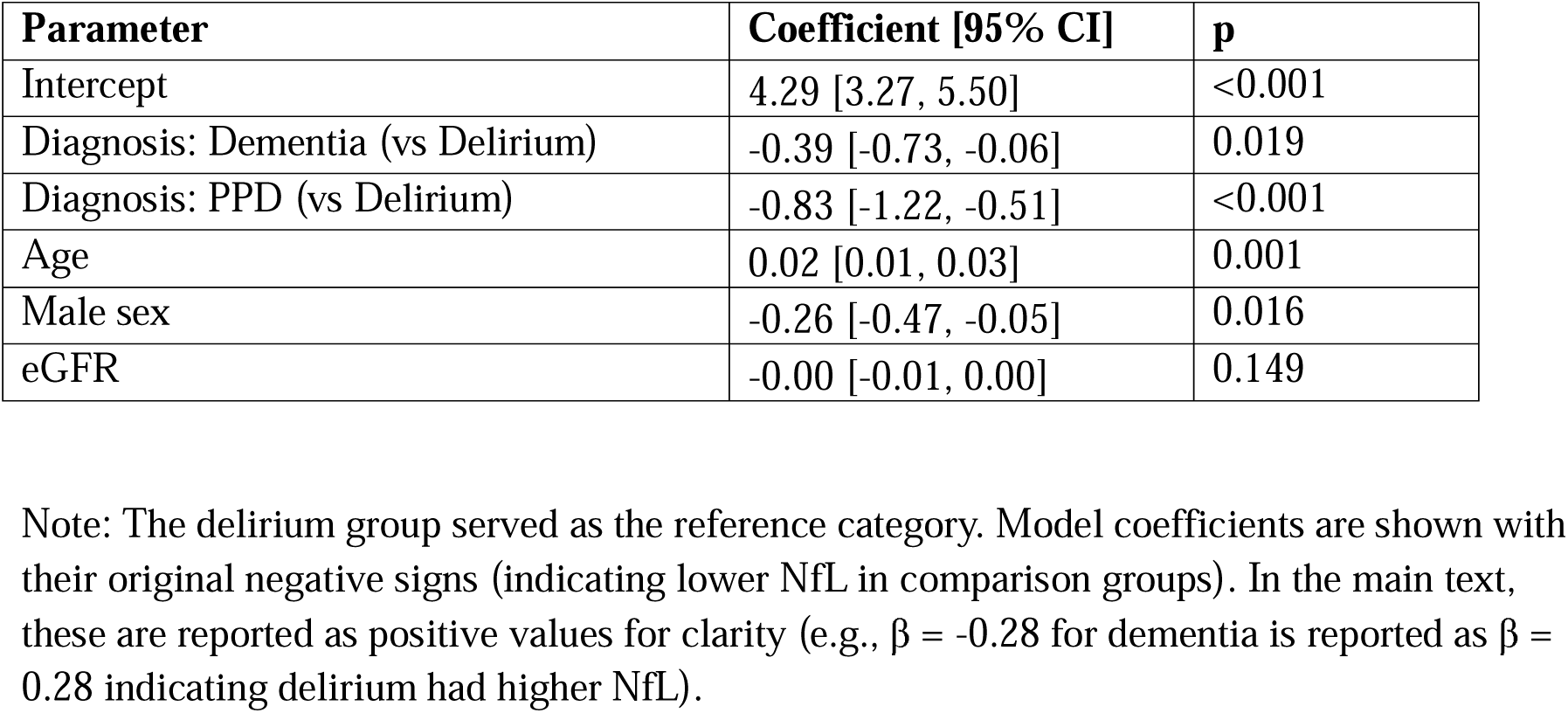
GFAP GLM (Reference = delirium) within Delirium Discovery Cohort.

**S3.** Plasma NfL x subgroup analysis of excluding primary psychiatric disorders (Reference = Delirium-only)

| **Parameter** | **Coefficients [95% CI]** | **p** |
| --- | --- | --- |
| Intercept | 4.78 [3.29, 6.30] | <0.001 |
| Diagnosis: Delirium + dementia (vs Delirium-only) | 0.36 [0.05, 0.66] | 0.019 |
| Diagnosis: Dementia (vs Delirium-only) | 1.05 [0.61, 1.52] | <0.001 |
| Age | 0.00 [-0.01, 0.02] | 0.568 |
| Male sex | 0.02 [-0.27, 0.30] | 0.882 |
| eGFR | -0.01 [-0.02, -0.01] | <0.001 |

**S4.**
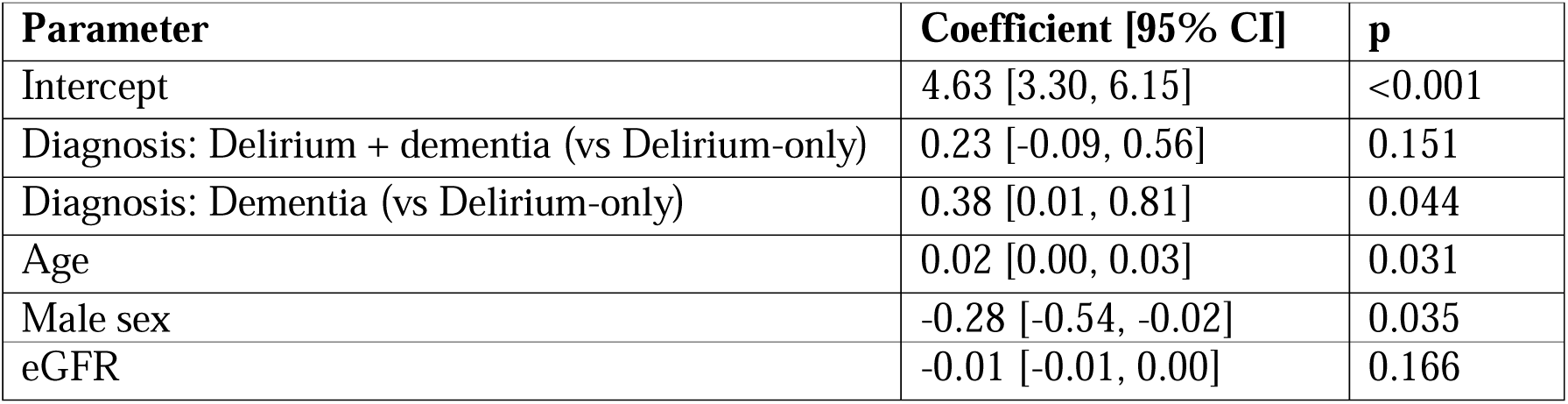
Plasma GFAP x subgroup analysis of excluding primary psychiatric disorders (Reference = Delirium-only)

| **Parameter** | **Coefficient [95% CI]** | **p** |
| --- | --- | --- |
| Intercept | 4.63 [3.30, 6.15] | <0.001 |
| Diagnosis: Delirium + dementia (vs Delirium-only) | 0.23 [-0.09, 0.56] | 0.151 |
| Diagnosis: Dementia (vs Delirium-only) | 0.38 [0.01, 0.81] | 0.044 |
| Age | 0.02 [0.00, 0.03] | 0.031 |
| Male sex | -0.28 [-0.54, -0.02] | 0.035 |
| eGFR | -0.01 [-0.01, 0.00] | 0.166 |

